# Cardiovascular Health Effects and Synthetic Cooling Agents in E-cigarettes Labeled as ‘clear’ Marketed in Massachusetts After the Tobacco Product Flavoring Ban

**DOI:** 10.1101/2024.04.18.24305863

**Authors:** Erika T. Minetti, Hanno C. Erythropel, Rachel Keith, Danielle R. Davis, Julie B. Zimmerman, Suchitra Krishnan-Sarin, Naomi M. Hamburg

## Abstract

**Introduction:** Massachusetts (MA) enacted statewide regulation on all flavored tobacco products in June 2020. Thereafter, electronic cigarettes (e-cigarettes) labeled ‘clear’ emerged on the market. We aimed to combine cardiovascular health effects with chemical analysis of ‘clear’ e-cigarettes.

**Methods:** We measured acute changes in blood pressure and heart rate following a 10-minute structured use of participants’ own e-cigarette, comparing ‘clear’ e-cigarette users with other flavored e-cigarette users and non-users. Chemical characterization and quantification of relevant flavorings and cooling agents (WS-3, WS-23) of 19 ‘clear’-labeled disposable e-cigarette liquids was carried out by GC/MS.

**Results:** After the ban, participants that used ‘clear’ labeled e-cigarettes increased from 0% to 21%. Increase in diastolic blood pressure and heart rate was significantly greater in ‘clear’ e-cigarettes users (n=22) compared to both non-’clear’ flavored e-cigarette users (n=114) and non-users (n=72). We saw similar results in heart rate when comparing Juul e-cigarette and ‘clear’ users; Juul was used as a reference as synthetic coolants WS-3 or WS-23 were not detected in these.

All (19/19) ‘clear’ e-liquids were found to contain synthetic cooling agents WS-23 and/or WS-3, menthol (18/19), as well as other flavorings (12/19).

**Discussion:** The detected presence of menthol alongside other flavorings in tested ‘clear’ products is a direct violation of the MA flavored tobacco product regulation, warranting stricter monitoring for new products and constituents. ‘clear’ e-cigarette use led to greater hemodynamic effects compared to other flavored e-cigarettes and Juul, which raises questions about the effect of cooling agents on users.

## Introduction

Flavored electronic cigarettes (e-cigarettes) appeal to youth/young adults and restricting access is a regulatory strategy to combat use. In June 2020, Massachusetts (MA) enacted a statewide regulation on “any flavored tobacco products,”^1^ specifically prohibiting the sale of products with “characterizing flavor” and only exempts “tobacco taste or aroma.” The introduction of similar regulation in California led to the emergence of tobacco products that contain novel ingredients, such as odorless synthetic coolants,^2^ which is currently under litigation.

After the MA flavor ban was enacted, e-cigarettes labeled as ‘clear’ became available. Though not always explicitly marketed as unflavored, the ‘clear’ labeling might suggest as such, and there have been limited investigations regarding composition and toxicity. We present the first study that combined untargeted chemical characterization to determine flavoring, as well as menthol and synthetic cooling agent presence in ‘clear’ e-liquids with examinations of the hemodynamic effects of acute exposure to ‘clear’ e-cigarettes.

## Methods

The Cardiovascular Injury due to Tobacco Use (CITU) study is an ongoing observational cohort study evaluating the health effects of e-cigarettes in young adults. Blood pressure (BP) and heart rate (HR) were examined in healthy young adults (ages 18-45) before and after a 10-minute structured use of participants’ own e-cigarette product or breathing through a straw (non-user controls) in the morning following abstinence from food, caffeine (>8 hours), and from tobacco and exercise (>6 hours).^3^ Changes in BP and HR measures were compared between users of ‘clear’ e-cigarettes (n=22), other non-’clear’ flavored e-cigarettes (n=114), and non-users (n=72).

For chemical analysis, 19 ‘clear’ disposables devices were purchased based on an online keyword search for ‘clear disposable’, including three brands reported by CITU participants (Crave, Luto, Space) between 4/2024-5/2025, and Juul ‘Menthol’ and ‘Virginia Tobacco’ in March 2024. E-liquid from each device was diluted and injected into a GC/MS for untargeted characterization, and into a GC/FID for quantification using commercially available standards following an established protocol.^4^

## Results

Cohort data was collected 04/2019-05/2023: ‘clear’ users: n=22, age 21±3, 46% female, 68% sole e-cigarette use; non-’clear’ flavored users: n=114, age 22±4, 45% female, 73% sole e-cigarette use; non-users: n=72, age 26±6, 54% female. Pre-ban, no ‘clear’ use was reported, whereas post-ban, 21% of participants used ‘clear’ and 79% used non-’clear’ flavored products, consistent with ongoing access to flavored products. Pre-use BP and HR did not differ between groups. Following acute exposure, use of ‘clear’ e-cigarettes resulted in a greater increase in diastolic BP and HR compared to both non-’clear’ flavored e-cigarette use and non-use, with a trend towards a greater increase in systolic BP (Figure 1A). In a sensitivity analysis restricted to sole e-cigarette users, the findings were similar.

**Figure:**
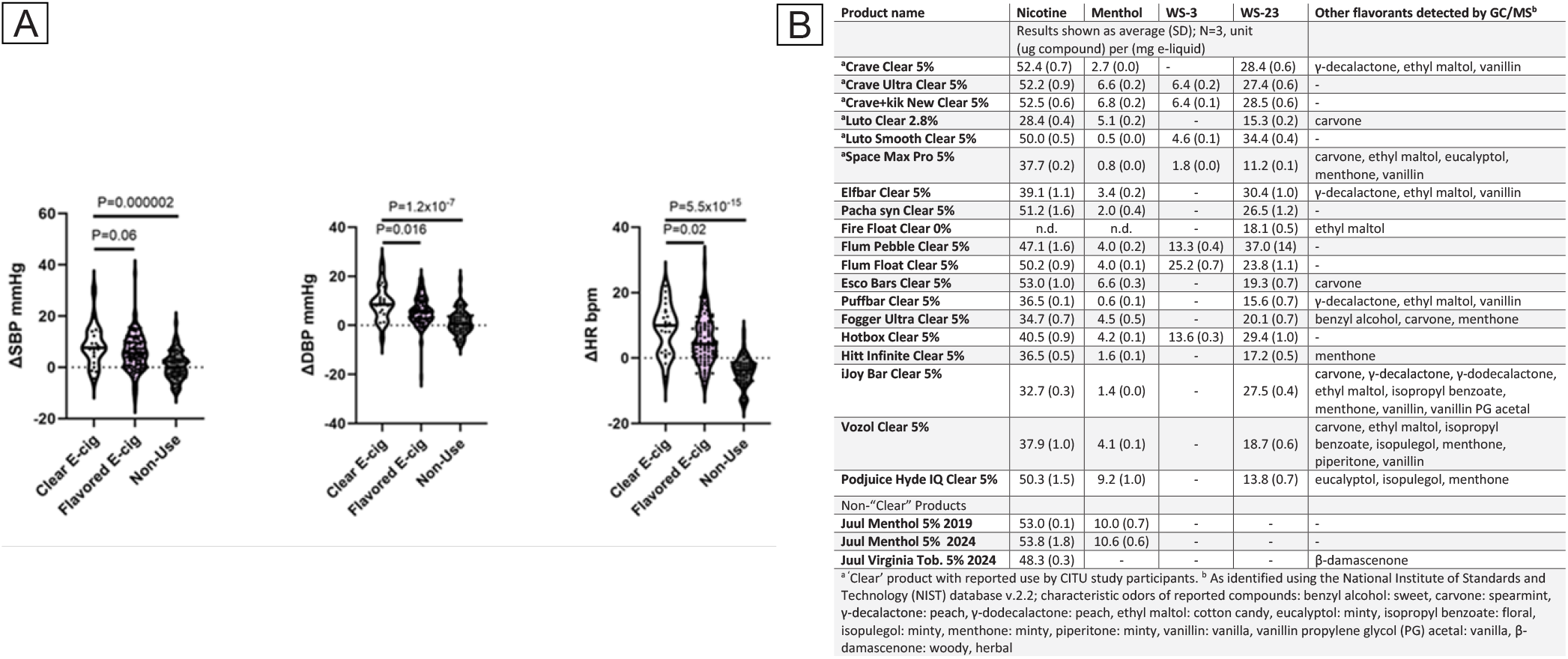
(**A**) Acute effects of ‘clear’ e-cigarettes on Blood Pressure (systolic: SBP; diastolic: DBP)) and Heart Rate (HR). Change in SBP, DBP, and HR differed across the three groups of users: ‘clear’ labeled e-cigarettes, flavored e-cigarette, and non-use by one way ANOVA (all P<0.001), post-hoc comparisons using LSD between ‘clear’ e-cig and other groups as noted. Statistical analyses were performed using SPSSv.27. (**B**) Disposable ‘clear’ and non-’clear’ products tested, quantified contents for nicotine, menthol, synthetic cooling agents WS-3 and WS-23, and a list of other flavorants detected by non-targeted GC/MS analysis.

Chemical analysis of ‘clear’ e-liquids demonstrated the presence of synthetic cooling agents WS-23 (19/19) and WS-3 (7/19), and menthol (18/19). In 12/19 products, other flavorings including menthone (peppermint flavor) carvone (spearmint), vanillin (vanilla), ethyl maltol (cotton candy), and γ-decalactone (peach) were detected (Figure 1B).

JUUL was the most common non-’clear’ flavored product (n=43/114) and no synthetic coolants were detected among JUUL products (Figure 1B). In a secondary analysis comparing JUUL and ‘clear’ exposure, we observed that post-use, increases in heart rate remained higher in ‘clear’ users compared to JUUL users (8.9±7.4 vs 4.5±8.1, P=0.01), whereas the change in diastolic BP was not significantly different (8.9±7.1 vs 6.4±6.0, P=0.1).

## Discussion

This study demonstrates that ‘clear’ e-cigarettes, which emerged following a tobacco product flavor ban in MA, contain significant levels of both menthol and synthetic, odorless cooling agents (WS-3, WS-23), and other flavorings. Exposure to ‘clear’ e-liquids, when compared with non-’clear’ flavored e-liquids, induced greater increases in BP and HR, including increased HR when compared with JUUL products that did not contain synthetic coolants.

The detected presence of menthol alongside other flavorings in tested ‘clear’ products is a direct violation of the MA flavored tobacco product regulation. Further, synthetic coolants (which are not specifically mentioned in the MA law) may reduce tobacco/nicotine harshness, facilitate deeper inhalation, and enhance nicotine delivery,^5^ which could help explain our hemodynamic findings and warrant their inclusion in tobacco flavor regulations. Notably, other countries have implemented restrictions on synthetic cooling agent use in combustible cigarettes.^4^

In conclusion, following the flavor ban in MA we observed the emergent use of new products by young adults that not only contained traditional and novel synthetic and odorless coolants, as well as other flavorings, but also produced differential hemodynamic effects when compared to other existing e-cigarettes. While this study is limited by chemical characterization of only ‘clear’ and JUUL e-liquids, and exposure only to the participants’ own products, our findings suggest that monitoring for new products and constituents including scientific evidence on their physiological and behavioral impacts are critically needed to maximize the impact of flavor bans and protect youth/young adults.

## Data Availability

All data produced in the present study are available upon reasonable request to the authors

## Disclaimer

The content is solely the responsibility of the authors and does not necessarily represent the views of the NIH, AHA or the FDA.

## Sources

1. 2019 Tobacco Control Law | Mass.gov. Accessed December 19, 2023. https://www.mass.gov/guides/2019-tobacco-control-law

2. Jabba S V., Erythropel HC, Anastas PT, Zimmerman JB, Jordt SE. Synthetic Cooling Agent and Other Flavor Additives in “Non-Menthol” Cigarettes Marketed in California and Massachusetts After Menthol Cigarette Bans. JAMA. 2023;330(17):1689–1691. doi:10.1001/JAMA.2023.17134

3. Keith RJ, Fetterman JL, Riggs DW, et al. Protocol to assess the impact of tobacco-induced volatile organic compounds on cardiovascular risk in a cross-sectional cohort: Cardiovascular Injury due to Tobacco Use study. BMJ Open. 2018;8(3):e019850. doi:10.1136/BMJOPEN-2017-019850

4. Jabba S V., Erythropel HC, Torres DG, et al. Synthetic Cooling Agents in US-marketed E-cigarette Refill Liquids and Popular Disposable E-cigarettes: Chemical Analysis and Risk Assessment. Nicotine Tob Res. 2022;24(7):1037–1046. doi:10.1093/NTR/NTAC046

5. Siddiqi TJ, Rashid AM, Siddiqi AK, et al. Association of Electronic Cigarette Exposure on Cardiovascular Health: A Systematic Review and Meta-Analysis. Curr Probl Cardiol. 2023;48(9). doi:10.1016/J.CPCARDIOL.2023.101748

